# Association between Hydroxyzine Use and Reduced Mortality in Patients Hospitalized for Coronavirus Disease 2019: Results from a multicenter observational study

**DOI:** 10.1101/2020.10.23.20154302

**Authors:** Nicolas Hoertel, Marina Sánchez, Raphaël Vernet, Nathanaël Beeker, Antoine Neuraz, Carlos Blanco, Mark Olfson, Cédric Lemogne, Pierre Meneton, Christel Daniel, Nicolas Paris, Alexandre Gramfort, Guillaume Lemaitre, Elisa Salamanca, Mélodie Bernaux, Ali Bellamine, Anita Burgun, Frédéric Limosin, On behalf of AP-HP / Universities / INSERM Covid-19 research collaboration and AP-HP Covid CDR Initiative

**Author notes:** Corresponding author Nicolas Hoertel, M.D., M.P.H., Ph.D., Corentin Celton Hospital, AP-HP.Centre, Paris University, 4 parvis Corentin Celton; 92130 Issy-les-Moulineaux, France, /.

## Abstract

**Objective:** To examine the association between hydroxyzine use and mortality in patients hospitalized for COVID-19, based on its anti-inflammatory and antiviral properties.

**Design:** Multicenter observational retrospective cohort study.

**Setting:** Greater Paris University hospitals, France.

**Participants:** 7,345 adults hospitalized for COVID-19 between 24 January and 1 April 2020, including 138 patients (1.9%) who received hydroxyzine during the visit at a mean dose of 49.8 mg (SD=51.5) for an average of 22.4 days (SD=25.9).

**Data source:** Assistance Publique-Hôpitaux de Paris Health Data Warehouse.

**Main outcome measures:** The study endpoint was death. We compared this endpoint between patients who received hydroxyzine and those who did not in time-to-event analyses adjusting for patient characteristics (such as age, sex, and comorbidities), clinical and biological markers of disease’s severity, and use of other medications. The primary analysis was a multivariable Cox model with inverse probability weighting. Sensitivity analyses included a multivariable Cox model and a univariate Cox regression model in a matched analytic sample in a 1:1 ratio.

**Results:** Over a mean follow-up of 20.3 days (SD=27.5), 994 patients (13.5%) had a primary end-point event. The primary multivariable analysis with inverse probability weighting showed a significant association between hydroxyzine use and reduced mortality (HR, 0.42; 95% CI, 0.25 to 0.71; p=0.001) with a significant dose-effect relationship (HR, 0.10; 95% CI, 0.02 to 0.45; p=0.003). This association was similar in sensitivity analyses. In secondary analyses conducted among subsamples of patients, we found a significant association between hydroxyzine use and a faster decrease in biological inflammatory markers associated with COVID-19-related mortality, including neutrophil-to-lymphocyte ratio (NLR), lymphocyte-to-C-reactive protein ratio (LCRP), and circulating interleukin 6 levels (IL-6) (all p<0.016), with a significant dose-effect relationship for NLR and LCRP (both p<0.037).

**Conclusions:** In this retrospective observational study, hydroxyzine use was associated with reduced mortality in patients hospitalized for COVID-19. This association may be partially mediated by specific anti-inflammatory properties of H1 antihistamines. Double-blind controlled randomized clinical trials of hydroxyzine for COVID-19 are needed to confirm these results.

## 1. Introduction

Global spread of the novel coronavirus SARS-CoV-2, the causative agent of coronavirus disease 2019 (COVID-19), has created an unprecedented infectious disease crisis worldwide.^1^ In the current absence of a vaccine or curative treatment, the search for an effective treatment for patients with COVID-19 among all available medications is urgently needed.^2 3^

Antihistamines are widely used in the treatment of urticaria, allergic rhinitis, hay fever, conjunctivitis and pruritus. They work by competitive binding to H1 receptors and inhibiting the action of histamine, a primary mediator of an early-phase allergic inflammation response that also modulates the late-phase response characterized by cellular influx of eosinophils, neutrophils, basophils, mononuclear cells, and T lymphocytes.^4^ *In vivo* and *in vitro* studies have also suggested additional anti-inflammatory properties of H1 antihistamines, including both receptor-dependent and receptor-independent mechanisms.^5^ The receptor-dependent mechanisms may involve inhibition of NF-kB dependent cytokines (such as IL-1, IL-2, IL-6, IL-8, IL-12, TNF-α)^6^ and adhesion proteins (such as ICAM-1, VCAM-1 and ECAM-1).^5^ The receptor-independent mechanisms, which require higher drug concentrations, may include the inhibition of the release by inflammatory cells of pre-formed mediators, such as histamine and eosinophil proteins, as well as eicosanoid generation and oxygen free radicals production.^5^

Prior research also supports *in vitro* antiviral activity of the H1 antihistamine hydroxyzine against MERS and hepatitis C virus,^7^ although no study to our knowledge has specifically studied its antiviral effect on SARS-CoV-2.

Among first generation antihistamines, hydroxyzine is one of the most prescribed antihistamines in France. Beyond its antihistaminic activity, hydroxyzine is also prescribed as a psychotropic medication for its tranquilizer and sedative properties, as it is a weak antagonist of the serotonin 5-HT2A, dopamine D2, and α1-adrenergic receptors.

Because prior research supports that severe COVID-19 is characterized by an excessive inflammatory response^8^ and that viral load could be associated with the worsening of symptoms,^9^ we hypothesized that hydroxyzine could be effective in reducing mortality among patients with COVID-19. Short-term use of hydroxyzine is generally well tolerated, although common side effects include sleepiness, headache, and dry mouth, and serious, less common serious ones may comprise delirium, QT prolongation, and torsade de pointes, particularly among older adults.^1^

Observational studies of patients with COVID-19 taking medications for other indications can help determine their efficacy for COVID-19, decide which should be prioritized for randomized clinical trials, and minimize the risk of patient exposure to potentially harmful and ineffective treatments. To our knowledge, no previous study has examined the potential efficacy of hydroxyzine for COVID-19.

For this purpose, we drew on data from the Assistance Publique-Hôpitaux de Paris (AP-HP) Health Data Warehouse, which includes data on all patients who have been admitted for COVID-19 to any of the 39 Greater Paris University hospitals.

In this report, we examined the association between hydroxyzine use and mortality among adult patients who have been admitted to these medical centers for COVID-19, using time-to-event analyses adjusting for potential confounders, including patient characteristics (sex, age, obesity, current smoking status, number of medical conditions associated with increased COVID-19-related mortality, any medication prescribed according to compassionate use or as part of a clinical trial, and the presence of current sleep or anxiety disorders), disease severity at hospital admission (using markers of clinical and biological severity of COVID-19), and other psychotropic medications that may influence disease prognosis,^10^ including any benzodiazepine or Z-drug, selective serotonin reuptake inhibitors (SSRIs) or serotonin and norepinephrine reuptake inhibitors (SNRIs), other antidepressants, mood stabilizers, and antipsychotic medications. We hypothesized that among patients hospitalized for COVID-19, hydroxyzine use would be independently associated with reduced mortality. We further hypothesized that this association would be mediated by a significantly faster decrease in biological inflammatory markers associated with COVID-19-related mortality among patients who used hydroxyzine during the visit compared to those who did not, as measured by high neutrophil-to-lymphocyte ratio (NLR), low lymphocyte-to-C-reactive protein ratio (LCRP), high circulating interleukin 6 levels (IL-6), and high lactate levels.^11-13^

## 2. Methods

### 2.1. Setting

We conducted this study at AP*-*HP, which includes 39 hospitals of which 23 are acute, 20 are adult and 3 are pediatric hospitals. We included all adults aged 18 years or over who have been admitted with COVID-19 to these medical centers from the beginning of the epidemic in France, i.e. January 24^th^, until April 1^st^. COVID-19 was ascertained by a positive reverse-transcriptase–polymerase-chain-reaction (RT-PCR) test from analysis of nasopharyngeal or oropharyngeal swab specimens. This observational study using routinely collected data received approval from the Institutional Review Board of the AP-HP clinical data warehouse (decision CSE-20-20_COVID19, IRB00011591). AP-HP clinical Data Warehouse initiative ensures patients’ information and consent regarding the different approved studies and data opt-out service through a transparency portal in accordance with European Regulation on data protection and authorization n°1980120 from National Commission for Information Technology and Civil Liberties (CNIL).

### 2.2. Data sources

We used data from the AP-HP Health Data Warehouse (‘Entrepôt de Données de Santé (EDS)’). This warehouse contains all the clinical data available on all inpatient visits for COVID-19 to any of the 39 Greater Paris University hospitals. The data obtained included patients’ demographic characteristics, vital signs, laboratory test and RT-PCR test results, medication administration data, current medication lists, current diagnoses, and death certificates.

### 2.3. Variables assessed

We obtained the following data for each patient at the time of the hospital admission: sex; age, which was categorized based on the OpenSAFELY study results^14^ (i.e. 18-50, 51-70, 71-80, 81+); obesity, defined as having a body-mass index higher than 30.0 kg/m^2^ or an International Statistical Classification of Diseases and Related Health Problems (ICD-10) diagnosis code for obesity (E66.0, E66.1, E66.2, E66.8, E66.9); self-reported current smoking status; the number of medical conditions associated with increased COVID-19-related mortality^14-17^ (categorized into 0, 1 and 2 or more conditions) based on ICD-10 diagnosis codes, including diabetes mellitus (E11), diseases of the circulatory system (I00-I99), diseases of the respiratory system (J00-J99), neoplasms (C00-D49), diseases of the blood and blood-forming organs and certain disorders involving the immune mechanism (D5-D8), delirium (F05, R41), and dementia (G30, G31, F01-F03); any sleep or anxiety disorder (G47*, F40-F48); any medication prescribed according to compassionate use or as part of a clinical trial (e.g. hydroxychloroquine, azithromycin, remdesivir, tocilizumab, sarilumab or dexamethasone); clinical severity of COVID-19 at admission, defined as having at least one of the following criteria:^11^ respiratory rate > 24 breaths/min or < 12 breaths/min, resting peripheral capillary oxygen saturation in ambient air < 90%, temperature > 40°C, or systolic blood pressure < 100 mm Hg; and biological severity of COVID-19 at admission, defined as having at least one of the following criteria:^11 12^ high neutrophil-to-lymphocyte ratio (NLR) or low lymphocyte-to-C-reactive protein ratio (LCR) (both variables were dichotomized at the median of the values observed values in the full sample; for NLR, the number of neutrophils and lymphocytes was taken per 10^9^/L, while LCR was calculated as lymphocyte count ((number/μL)/C-reactive protein (mg/dL)), or plasma lactate levels higher than 2 mmol/L; any other antihistamine medications; any prescribed psychotropic medication, including any benzodiazepine or Z-drug, any SSRI or SNRI, any other antidepressant, any mood stabilizer (i.e. lithium or antiepileptic medications with mood stabilizing effects), and any antipsychotic medication.

All medical notes and prescriptions are computerized in Greater Paris University hospitals. Medications and their mode of administration (i.e., dosage, frequency, date, condition of intake) were identified from medication administration data or scanned hand-written medical prescriptions, through two deep learning models based on BERT contextual embeddings,^18^ one for the medications and another for their mode of administration. The model was trained on the APmed corpus,^19^ a previously annotated dataset for this task. Extracted medications names were then normalized to the Anatomical Therapeutic Chemical (ATC) terminology using approximate string matching.

### 2.4. Hydroxyzine use

Study baseline was defined as the date of hospital admission. Hydroxyzine use was defined as receiving this medication at any time during the follow-up period, from study baseline to the end of the index hospitalization or death.

### 2.5. Endpoint

The endpoint was the time from study baseline to death. Patients without an end-point event had their data censored on April 1^st^, 2020.

### 2.6. Statistical analysis

We calculated frequencies and means (± standard deviations (SD)) of each baseline characteristic described above in patients receiving or not receiving hydroxyzine and examined their potential between-group imbalance as shown by standardized mean differences higher than 0.1.^20^

To examine the associations between hydroxyzine use and the endpoint, we performed Cox proportional-hazard regression models. To help account for the nonrandomized prescription of hydroxyzine and reduce the effects of confounding, the primary analysis used propensity score analysis with inverse probability weighting.^21 22^ The individual propensities for hydroxyzine use were estimated using a multivariable logistic regression model that included patients’ characteristics (sex, age, obesity, current smoking status, number of medical conditions associated with increased COVID-19-related mortality, the presence of current sleep or anxiety disorder, any medication prescribed according to compassionate use or as part of a clinical trial), disease’s severity at hospital admission (using markers of clinical and biological severity of COVID-19), and any other antihistamine medication and any psychotropic medications (any benzodiazepine or Z-drug, any SSRI or SNRI antidepressant, any other antidepressant, any mood stabilizer, and any antipsychotic medication). In the inverse-probability-weighted analyses, the predicted probabilities from the propensity-score models were used to calculate the stabilized inverse-probability-weighting weights.^21^ Associations between hydroxyzine use and the endpoint was then estimated using a multivariable Cox regression model including the inverse-probability-weighting weights. Kaplan-Meier curves were performed using the inverse-probability-weighting weights,^23^ and their pointwise 95% confidence intervals were estimated using the nonparametric bootstrap method.^24^

We conducted two sensitivity analyses, including a multivariable Cox regression model with the same covariates as the inverse-probability-weighted analyses, and a univariate Cox regression model in a matched analytic sample using a 1:1 ratio, based on the same variables used for both the inverse-probability-weighted and the multivariable Cox regression analyses. In that latter analysis, to reduce the effects of confounding, optimal matching was used to obtain the smallest average absolute distance across all clinical characteristics between exposed patient and non-exposed matched controls. Weighted Cox regression models were used when proportional hazards assumption was not met.

We also performed four additional analyses. First, to increase our confidence that the results might not be due to unmeasured confounding or indication bias, we examined (i) this association among patients who received hydroxyzine only within the 3 months before hospital admission as compared to those who received it during the visit only, and (ii) the change of the magnitude of the effect of potential residual confounding on our results by varying the relationship of each potential confounder with the endpoint.^25^ Second, to examine a potential immortal bias in the exposed group (null probability of dying during the period between study baseline and the initiation of hydroxyzine), we performed additional Cox proportional-hazard regression analyses to compare the potential effect of hydroxyzine use to that of an active comparator, i.e., zopiclone. We chose this comparator because most hydroxyzine-treated participants (59.4%) received hydroxyzine at low doses (i.e., 25 mg per day or less) for sleep problems.^26^ Third, we examined a potential dose-effect relationship by testing the association between the daily dose received (dichotomized by the median dose) with the endpoint within patients receiving hydroxyzine. Finally, if a significant association were found between hydroxyzine use during the visit and mortality, we planned to examine whether this association could be at least partially explained by a significantly greater decrease in biological inflammatory markers associated with increased COVID-19-related mortality, as measured by high neutrophil-to-lymphocyte ratio, low lymphocyte-to-C-reactive protein, high circulating interleukin 6 levels, and high lactate levels, among patients who used hydroxyzine during the visit compared to those who did not. To this end, we performed two-way repeated measures ANOVA among the subsamples of patients for whom each biological inflammatory marker was assessed at least twice during the visit. For patients with three or more measures, we only considered the first and the last measure of the visit. Log transformation was used to address the non-normal distribution of the biological markers. We also searched for a potential dose-effect relationship by testing the association between the daily dose received (dichotomized by the median dose) with changes in the biological markers between the first and the last measure of the visit.

For all associations, we performed residual analyses to assess the fit of the data, check assumptions, including proportional hazards assumptions, and examined the potential influence of outliers. To improve the quality of result reporting, we followed the recommendations of The Strengthening the Reporting of Observational Studies in Epidemiology (STROBE) Initiative.^27^ Statistical significance was fixed *a priori* at two-sided p-value<0.05. All analyses were conducted in R software version 2.4.3 (R Project for Statistical Computing).

## 3. Results

### 3.1. Characteristics of the cohort

Of the 9,509 patients with a positive COVID-19 RT-PCR test consecutively admitted to the hospital, a total of 2,164 patients (22.8%) were excluded because of missing data or their young age (i.e. less than 18 years old of age). Of the remaining 7,345 adult patients, 138 (1.9%) patients received hydroxyzine during the hospitalization, for an average of 22.4 days (SD=25.9, median=12.5 days, minimum=1 day, maximum=114 days), at a mean daily dose of 49.8 mg (SD=51.5, median=25 mg, minimum=12.5 mg, maximum=300.0 mg) (**Figure 1**). Mean time from study baseline to first hydroxyzine prescription was of 7.6 days (SD=10.4; median = 4 days; range: 0 to 63 days).

**Figure 1.**
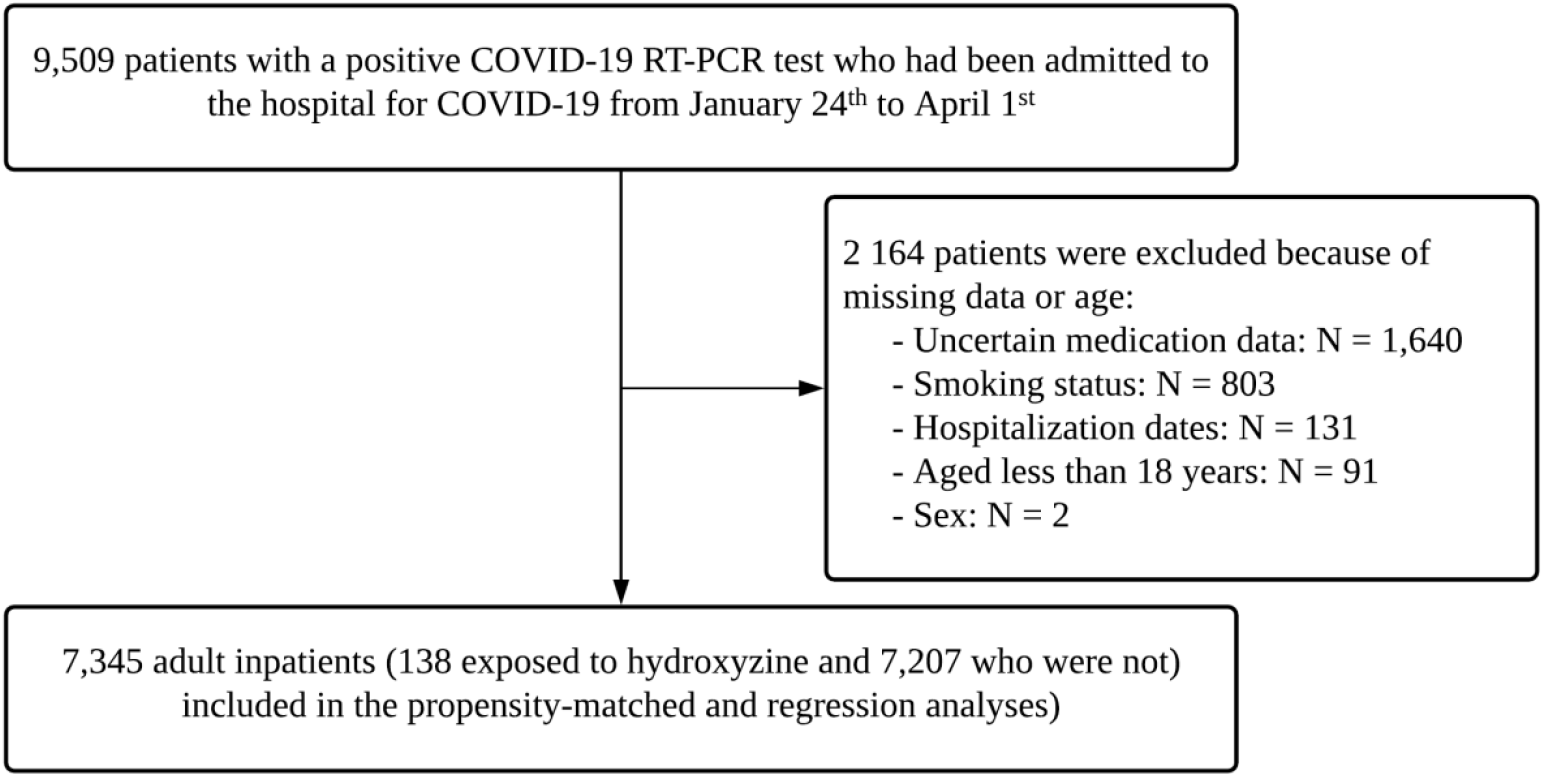
Study cohort.

COVID-19 RT-PCR test results were obtained after a mean delay of 5 days (SD=11.7, median=1 day) from the date of hospital admission. This delay was not significantly different between patients receiving or not receiving hydroxyzine [mean delay in the exposed group=7.1 day (SD=14.9); mean delay in the non-exposed group=5.0 day (SD=11.7); Welch’s t-test=-1.63, p=0.106)].

Over a mean follow-up of 20.3 days (SD=27.5; median=7 days; range: 1 day to 117 days), 994 patients (13.5%) had an end-point event at the time of data cutoff on April 1^st^. Among patients who received hydroxyzine, the mean follow-up was 22.4 days (SD=25.9; median=12.5 days; range: 1 day to 114 days), while it was of 20.2 days (SD=27.5; median=6 days; range: 4 day to 117 days) in those who did not (Welch’s t-test=-0.97, p=0.336). All baseline characteristics were significantly and independently associated with mortality, except for current smoking, any current sleep or anxiety disorder, any other antidepressant, any mood stabilizer, and any antipsychotic medication (**eTable 1**).

The distribution of patients’ characteristics according to hydroxyzine use is shown in **Table 1**. In the full sample, hydroxyzine use substantially differed according to all baseline characteristics (**Table 1**). The direction of these associations indicated older age and overall greater medical severity of patients receiving hydroxyzine than those who did not. After applying the propensity score weights and in the matched analytic sample, these differences were substantially reduced (**Table 1**).

**Table 1.** Characteristics of patients with COVID-19 receiving or not receiving hydroxyzine.

|  | Exposed to<br>hydroxyzine<br>(N=138) | Not exposed to<br>hydroxyzine<br>(N=7,207) | Non-exposed<br>matched group<br>(N=138) | Exposed to<br>hydroxyzine<br>vs.<br>Not exposed to<br>hydroxyzine | Exposed to hydroxyzine<br>vs.<br>Not exposed to<br>hydroxyzine | Exposed to<br>hydroxyzine<br>vs.<br>Non-exposed<br>matched group |
| --- | --- | --- | --- | --- | --- | --- |
|  |  |  |  | Standardized mean<br>differences | Weighted standardized<br>mean differences | Standardized mean<br>differences in the<br>matched analytic<br>sample |
|  | N (%) | N (%) | N (%) | SMD | SMD | SMD |
| Age |  |  |  | 0.322* | 0.057 | 0.162* |
| 18 to 50 years | 35 (25.4%) | 2674 (37.1%) | 26 (18.8%) |  |  |  |
| 51 to 70 years | 59 (42.8%) | 2471 (34.3%) | 64 (46.4%) |  |  |  |
| 71 to 80 years | 27 (19.6%) | 915 (12.7%) | 28 (20.3%) |  |  |  |
| More than 80 years | 17 (12.3%) | 1147 (15.9%) | 20 (14.5%) |  |  |  |
| Sex |  |  |  | 0.225* | 0.043 | 0.045 |
| Women | 53 (38.4%) | 3566 (49.5%) | 50 (36.2%) |  |  |  |
| Men | 85 (61.6%) | 3641 (50.5%) | 88 (63.8%) |  |  |  |
| Obesity <sup>a</sup> |  |  |  | 0.299* | 0.059 | 0.104* |
| Yes | 34 (24.6%) | 941 (13.1%) | 28 (20.3%) |  |  |  |
| No | 104 (75.4%) | 6266 (86.9%) | 110 (79.7%) |  |  |  |
| Smoking <sup>b</sup> |  |  |  | 0.254* | 0.046 | 0.081 |
| Yes | 23 (16.7%) | 600 (8.33%) | 19 (13.8%) |  |  |  |
| No | 115 (83.3%) | 6607 (91.7%) | 119 (86.2%) |  |  |  |
| Number of medical conditions <sup>γ</sup> |  |  |  | 0.797* | 0.161* | 0.080 |
| 0 | 40 (29.0%) | 4710 (65.4%) | 42 (30.4%) |  |  |  |
| 1 | 13 (9.42%) | 521 (7.23%) | 10 (7.3%) |  |  |  |
| 2 or more | 85 (61.6%) | 1976 (27.4%) | 86 (62.3%) |  |  |  |
| Medication according to<br>compassionate use or as part of<br>a clinical trial <sup>θ</sup> |  |  |  | 0.489* | 0.098 | 0.015 |
| Yes | 53 (38.4%) | 1235 (17.1%) | 52 (37.7%) |  |  |  |
| No | 85 (61.6%) | 5972 (82.9%) | 86 (62.3%) |  |  |  |
| Anxiety or insomnia <sup>ε</sup> |  |  |  | 0.333* | 0.101* | 0.044 |
| Yes | 18 (13.0%) | 281 (3.90%) | 16 (11.6%) |  |  |  |
| No | 120 (87.0%) | 6926 (96.1%) | 122 (88.4%) |  |  |  |
| Any other antihistamine<br>medication |  |  |  | 0.247* | 0.065 | <0.001 |
| Yes | 8 (5.8%) | 92 (1.3%) | 8 (5.8%) |  |  |  |
| No | 130 (94.2%) | 7115 (98.7%) | 130 (94.2%) |  |  |  |
| SSRI or SNRI |  |  |  | 0.299* | 0.039 | <0.001 |
| Yes | 17 (12.3%) | 301 (4.2%) | 17 (12.3%) |  |  |  |
| No | 121 (87.7%) | 6906 (95.8%) | 121 (87.7%) |  |  |  |
| Any other antidepressant |  |  |  | 0.175* | 0.030 | <0.001 |
| Yes | 7 (5.1%) | 135 (1.9%) | 7 (5.1%) |  |  |  |
| No | 131 (94.9%) | 7072 (98.1%) | 131 (94.9%) |  |  |  |
| Any mood stabilizer medication<br><sup>ω</sup> |  |  |  | 0.297* | 0.059 | <0.001 |
| Yes | 16 (11.6%) | 271 (3.76%) | 16 (11.6%) |  |  |  |
| No | 122 (88.4%) | 6936 (96.2%) | 122 (88.4%) |  |  |  |
| Any benzodiazepine or Z-drug |  |  |  | 0.564* | 0.140* | 0.016 |
| *Yes* | 44 (31.9%) | 708 (9.82%) | 43 (31.2%) |  |  |  |
| *No* | 94 (68.1%) | 6499 (90.2%) | 95 (68.8%) |  |  |  |
| Any antipsychotic |  |  |  | 0.455* | 0.134* | <0.001 |
| *Yes* | 23 (16.7%) | 241 (3.34%) | 23 (16.7%) |  |  |  |
| *No* | 115 (83.3%) | 6966 (96.7%) | 115 (83.3%) |  |  |  |
| Clinical severity of Covid-19 at admission <sup>μ</sup> |  |  |  | 0.635* | 0.120* | 0.030 |
| *Yes* | 53 (38.4%) | 1511 (21.0%) | 51 (37.0%) |  |  |  |
| *No* | 51 (37.0%) | 1807 (25.1%) | 52 (37.7%) |  |  |  |
| *Missing* | 34 (24.6%) | 3889 (54.0%) | 35 (25.4%) |  |  |  |
| Biological severity of Covid-19 at admission <sup>κ</sup> |  |  |  | 0.508* | 0.047 | 0.138* |
| *Yes* | 69 (50.0%) | 2370 (32.9%) | 75 (54.3%) |  |  |  |
| *No* | 42 (30.4%) | 1819 (25.2%) | 43 (31.2%) |  |  |  |
| *Missing* | 27 (19.6%) | 3018 (41.9%) | 20 (14.5%) |  |  |  |

<sup>α</sup> Defined as having a body-mass index higher than 30 kg/m<sup>2</sup> or an International Statistical Classification of Diseases and Related Health Problems (ICD-10) diagnosis code for obesity (E66.0, E66.1, E66.2, E66.8, E66.9).
<sup>β</sup> Smoking status was self-reported.
<sup>γ</sup> Assessed using ICD-10 diagnosis codes for diabetes mellitus (E11), diseases of the circulatory system (I00-I99), diseases of the respiratory system (J00-J99), neoplasms (C00-D49), diseases of the blood and blood-forming organs and certain disorders involving the immune mechanism (D5-D8), delirium (F05, R41) and dementia (G30, G31, F01-F03).
<sup>θ</sup> Any medication prescribed as part of a clinical trial or according to compassionate use (e.g., hydroxychloroquine, azithromycin, remdesivir, tocilizumab, sarilumab or dexamethasone).
<sup>ε</sup> Assessed using ICD-10 diagnosis codes for anxiety, dissociative, stress-related, somatoform and other nonpsychotic mental disorders (F40-F48) and insomnia (G47).
<sup>Ω</sup> Included lithium or antiepileptic medications with mood stabilizing properties.
<sup>u</sup> Defined as having at least one of the following criteria: respiratory rate > 24 breaths/min or < 12 breaths/min, resting peripheral capillary oxygen saturation in ambient air < 90%, temperature > 40°C, or systolic blood pressure < 100 mm Hg.
<sup>k</sup> Defined as having at least one of the following criteria: high neutrophil-to-lymphocyte ratio, low lymphocyte-to-C-reactive protein (both variables were dichotomized at the median of the values observed in the full sample), and plasma lactate levels higher than 2 mmol/L.
\* Mean standardized difference indicate substantial between-group imbalance ( $p > 0.1$ )
Abbreviations: HR, hazard ratio; SE, standard error.

### 3.2. Study endpoint

Among patients receiving hydroxyzine, death occurred in 15 patients (10.9%), while 979 non-exposed patients (16.6%) had this outcome (**Table 2**). Despite the older age and the overall greater medical severity of patients receiving hydroxyzine than those who did not, unadjusted hazard ratio of the association between hydroxyzine use and reduced mortality was close to statistical significance (HR, 0.60, 95% CI, 0.36 to 1.00, p=0.051) (**Table 2**). When taking into account differences in baseline characteristics between exposed and non-exposed patients, the primary multivariable analysis with inverse probability weighting showed a significant association between hydroxyzine use and mortality (HR, 0.42; 95% CI, 0.25 to 0.71, p=0.001) (**Table 2, Figure 2**).

**Table 2.** Association between hydroxyzine use and the endpoint of death in the full sample and in the matched analytic sample.

|  | Number of events /<br>Number of patients | Crude Cox<br>regression<br>analysis | Multivariable Cox<br>regression analysis<br>(df=21) | Analysis weighted by<br>inverse-probability-weighting weights | Number of events<br>/ Number of<br>patients | Univariate Cox<br>regression in the<br>matched analytic<br>sample (1:1) |
| --- | --- | --- | --- | --- | --- | --- |
|  | N (%) | HR (95% CI;<br>p-value) | HR (95% CI;<br>p-value)<sup>a</sup> | HR (95% CI;<br>p-value) | N (%) | HR (95% CI;<br>p-value) |
| Hydroxyzine | 15 / 138 (10.9%) | 0.60 (0.36 – 1.00;<br>0.051) | 0.42 (0.25 – 0.71;<br>0.001*) | 0.42 (0.25 – 0.71;<br>0.001*) | 15 / 138 (10.9%) | 0.39 (0.21 – 0.72;<br>0.003*) |
| No hydroxyzine | 979 / 7,207 (16.6%) | Ref. | Ref. | Ref. | 29 / 138 (21.0%) | Ref. |

\* p-value is significant (p<0.05).

**Figure 2.**
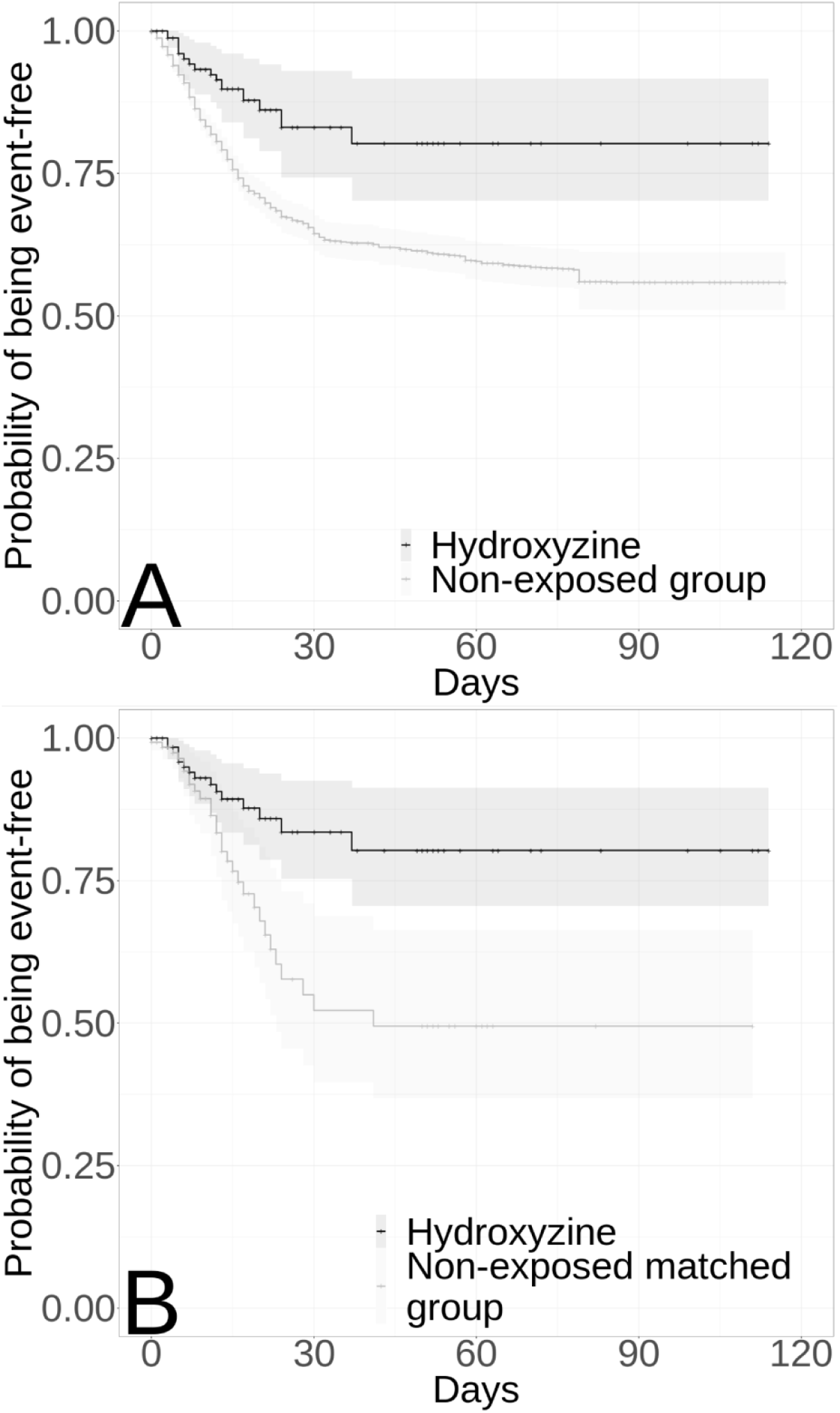
Kaplan-Meier curves for death in the full sample (A) (N=7,345) and in the matched analytic sample (B) (N=276) of patients who had been admitted to the hospital for Covid-19 according to hydroxyzine use. The shaded areas represent pointwise 95% confidence intervals.

In sensitivity analyses, the multivariable Cox regression model in the full sample yielded similar results (HR, 0.42; 95% CI, 0.24 to 0.71, p=0.001), as did the univariate Cox regression model in the matched analytic sample (HR, 0.39; 95% CI, 0.21 to 0.72 p=0.003) (**Table 2, Figure 2**).

Additional analyses indicated that patients who were prescribed hydroxyzine in the three months before the admission and not during the visit were significantly at higher risk of death than those who received this medication only during the visit (**eTable 2**).

The quantitative bias analysis with observed imbalances showed fairly robust hazard ratios under a wide range of assumed associations between potential confounders and the endpoint. Associations for an apparent hazard ratio of 0.42 are presented in **eFigure 2**. The required strength of the associations between each covariate and the outcome for the true hazard ratio to be substantially altered is considerably greater than those observed in the sample, suggesting that residual confounding is unlikely to affect our results.

When considering zopiclone as an active comparator, we found that hydroxyzine use was associated with reduced mortality in the crude (HR, 0.38; 95% CI, 0.21 to 0.70, p=0.002) and in the main (HR, 0.32; 95% CI, 0.15 to 0.70, p=0.004) analyses, as well as in sensitivity analyses (**eTable 3**). Zopiclone use was not associated with decreased mortality compared to participants not receiving either medication.

Finally, exposure to higher (median daily dose=75 mg, SD=63.6) rather than lower (median daily dose=25 mg, SD=4.2) doses of hydroxyzine were significantly associated with reduced mortality (**eTable 4**).

A *post-hoc* analysis indicated that in the full sample, we had 80% power to detect hazard ratios for hydroxyzine of at least 0.32 and 0.18 in the full sample and in the matched analytic sample, respectively.

### 3.3. Course of the levels of the biological inflammatory markers NLR, LCRP, IL-6, and lactates during the visit according to hydroxyzine use

The levels of NLR, LCRP, IL-6, and lactates were significantly associated with the endpoint of death in subsamples of patients for whom each marker was assessed at least twice during the visit (ranging from 315 for IL-6 to 2,158 for NLR) (**eTable 5**). Hydroxyzine use was significantly associated with changes between the first and last levels of LCRP, NLR, and IL-6 (**Figure 3; eTable 6**). It was associated with a 431.9% increase in the LCRP (versus a 95.9% increase in the non-exposed group, p<0.001), a 40.9% decrease in the NLR (versus a 4.2% decrease in the non-exposed group, p<0.001), and a 76.6% decrease in IL-6 (versus a 34.7% increase in the non-exposed group, p=0.016). The mean time between the first and last measure was 11.6 days (SD=11.8) for LCRP [mean=12.3 days, SD=8.7 in patients receiving hydroxyzine; mean=11.6 days, SD=11.9 in those who did not], 11.9 days (SD=12.5) for NLR [mean=13.4 days, SD=11.0 in patients receiving hydroxyzine; mean=11.9, SD=12.2 in those who did not], and 7.5 days (SD=7.1) for IL-6 (mean=7.4 days, SD=3.6 in patients receiving hydroxyzine; mean=7.5 days, SD=3.6 in those who did not). We found a significant dose-effect relationship between the daily dose received and the changes in the biological markers between the first and the last measure of the visit for NLR and LCRP. Exposure to higher *versus* lower doses of hydroxyzine was associated with a 382.6% increase in the LCRP (versus a 250.0% increase with lower doses, p=0.037) and a 63.5% decrease in the NLR (versus a 9.8% decrease with lower doses, p=0.021) (**eFigure 1; eTable 7**).

**Figure 3.**
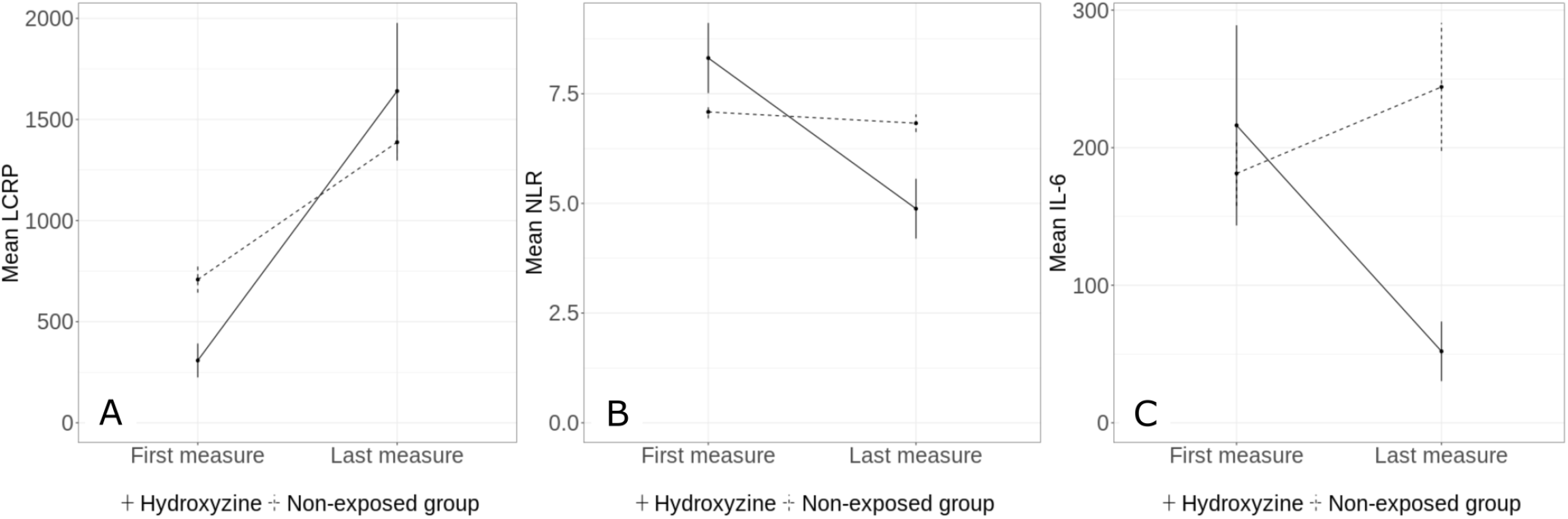
Course of the levels of the biological inflammatory markers LCRP (A), NLR (B) and IL-6 (C) during the visit between patients with COVID-19 receiving hydroxyzine and those who did not. Vertical lines represent standard errors. Note: For NLR, the number of neutrophils and lymphocytes was taken per 10^9^/L, while LCRP was calculated as follows: lymphocyte count (number/μL)/C-reactive protein (mg/dL). Abbreviations: LCRP, lymphocyte-to-C-reactive protein ratio; NLR, neutrophil-to-lymphocyte ratio: IL-6, circulating interleukin 6 levels.

## 4. Discussion

In this multicenter retrospective observational study involving a large sample of patients admitted to the hospital for COVID-19, our results suggest that hydroxyzine use, at a median daily dose of 25 mg (SD=51.5) for a median duration of 12.5 days (SD=25.9), was significantly and substantially associated with reduced mortality, independently of patients’ characteristics, clinical and biological markers of disease’s severity at hospital admission, and use of other medications. This association was significantly stronger at higher rather than lower doses of hydroxyzine. We also found among patients who used hydroxyzine during the admission compared to those who did not that this association was associated with a significantly faster decrease in biological inflammatory markers associated with increased COVID-19-related mortality, i.e., NLR, LCRP and IL-6. This association had a significant dose-effect relationship for NLR and LCRP. These findings should be interpreted with caution due to the observational design of the study. However, our findings provide support for conducting controlled randomized clinical trials of hydroxyzine for COVID-19.

Our study has several limitations. First, there are two possible major potential inherent biases in observational studies: unmeasured confounding and confounding by indication. We tried to minimize the effects of confounding in different ways. First, we used a multivariable regression model with inverse probability weighting to minimize the effects of confounding by indication.^21 22^ Second, we performed sensitivity analyses, including a multivariable Cox regression models and an univariate Cox regression model in a matched analytic sample, that showed similar results. Third, although some amount of unmeasured confounding may remain, our analyses adjusted for numerous potential confounders and a quantitative bias analysis with observed imbalances suggested that residual confounding is unlikely to affect our results. Fourth, the significant dose-effect relationship further supported our conclusion. Fifth, the use of an active comparator, zopiclone, yielded similar results. Finally, the association was only observed in patients who received hydroxyzine during the visit and not in those who received it only within the 3 months before hospital admission.

A second limitation includes missing data for some baseline characteristic variables, including baseline clinical and biological markers of severity of COVID-19, which may be explained by the overwhelming of all hospital units during the COVID-19 peak incidence, and potential for inaccuracies in the electronic health records in this context, especially for hand-written medical prescriptions. However, the associations observed between baseline characteristics and mortality are in line with prior epidemiological studies.^14^ Third, inflation of type I error might have occurred in secondary exploratory analyses due to multiple testing. Finally, despite the multicenter inpatient design, our results may not be generalizable to other settings or regions.

We found a significant association between hydroxyzine use and a faster decrease in biological inflammatory markers associated with COVID-19-related mortality, including NLR, LCRP, and IL-6, with a significant dose-effect relationship for NLR and LCRP. These findings suggest that the association between hydroxyzine use and reduced mortality might be at least partially explained by the anti-inflammatory properties of H1antihistamines. They may involve receptor-dependent mechanisms such as the possible inhibition of NF-kB dependent cytokines (such as IL-1, IL-2, IL-6, IL-8, IL-12, TNF-α)^6^ and adhesion proteins (such as ICAM-1, VCAM-1 and ECAM-1),^5^ and receptor-independent mechanisms such as the inhibition of the release by inflammatory cells of pre-formed mediators, such as histamine and eosinophil proteins, and eicosanoid generation and oxygen free radicals production.^5^

In this multicenter observational retrospective study involving patients admitted to the hospital for COVID-19, hydroxyzine use was significantly and substantially associated with reduced mortality, independently of background patient characteristics, clinical and biological markers of disease’s severity, and use of other medications, with a significant dose-effect relationship. This association might be explained by the known anti-inflammatory properties of H1 antihistamines as our results suggest a significant association between hydroxyzine use and a faster decrease in several biological inflammatory markers associated with COVID-19-related mortality, including NLR, LCRP, and IL-6, with a significant dose-effect relationship for NLR and LCRP. Double blind controlled randomized clinical trials of hydroxyzine and other antihistamines H1 for COVID-19 are needed to confirm these results.

## Data Availability

Data from the AP-HP Health Data Warehouse can be obtained at https://eds.aphp.fr//.

## Acknowledgments

The authors thank the EDS APHP COVID consortium integrating the APHP Health Data Warehouse team as well as all the APHP staff and volunteers who contributed to the implementation of the EDS-COVID database and operating solutions for this database. Collaborators of the EDS APHP COVID consortium are: Pierre-Yves ANCEL, Alain BAUCHET, Nathanaël BEEKER, Vincent BENOIT, Mélodie BERNAUX, Ali BELLAMINE, Romain BEY, Aurélie BOURMAUD, Stéphane BREANT, Anita BURGUN, Fabrice CARRAT, Charlotte CAUCHETEUX, Julien CHAMP, Sylvie CORMONT, Christel DANIEL, Julien DUBIEL, Catherine DUCLOAS, Loic ESTEVE, Marie FRANK, Nicolas GARCELON, Alexandre GRAMFORT, Nicolas GRIFFON, Olivier GRISEL, Martin GUILBAUD, Claire HASSEN-KHODJA, François HEMERY, Martin HILKA, Anne-Sophie JANNOT, Jerome LAMBERT, Richard LAYESE, Judith LEBLANC, Léo LEBOUTER, Guillaume LEMAITRE, Damien LEPROVOST, Ivan LERNER, Kankoe LEVI SALLAH, Aurélien MAIRE, Marie-France MAMZER, Patricia MARTEL, Arthur MENSCH, Thomas MOREAU, Antoine NEURAZ, Nina ORLOVA, Nicolas PARIS, Bastien RANCE, Hélène RAVERA, Antoine ROZES, Elisa SALAMANCA, Arnaud SANDRIN, Patricia SERRE, Xavier TANNIER, Jean-Marc TRELUYER, Damien VAN GYSEL, Gaël VAROQUAUX, Jill Jen VIE, Maxime WACK, Perceval WAJSBURT, Demian WASSERMANN, Eric ZAPLETAL.

## Contributors

NH designed the study, performed statistical analyses, and wrote the first draft of the manuscript. MSR contributed to study design, performed statistical analyses and critically revised the manuscript. FL contributed to study design and critically revised the manuscript for scientific content. RV contributed to statistical analyses and critically revised the manuscript for scientific content. NB and ASJ contributed to study design and critically revised the manuscript for scientific content. NB, ASJ, AN, NP, CD, AG, GL, MB, and AB contributed to database build process. Other authors critically revised the manuscript for important scientific content. NH is the guarantor. The corresponding author attests that all listed authors meet authorship criteria and that no others meeting the criteria have been omitted.

## Ethical approval

This observational study using routinely collected data received approval from the Institutional Review Board of the AP-HP clinical data warehouse (decision CSE-20-20_COVID19, IRB00011591). AP-HP clinical Data Warehouse initiative ensures patients’ information and informed consent regarding the different approved studies through a transparency portal in accordance with European Regulation on data protection and authorization n°1980120 from National Commission for Information Technology and Civil Liberties (CNIL). All procedures related to this work adhered to the ethical standards of the relevant national and institutional committees on human experimentation and with the Helsinki Declaration of 1975, as revised in 2008.

## Data sharing

Data from the AP-HP Health Data Warehouse can be obtained at https://eds.aphp.fr//.

<u>The corresponding author affirms that the manuscript is an honest, accurate, and transparent account of the study being reported. No important aspects of the study have been omitted and any discrepancies from the study as planned have been disclosed</u>.

## Dissemination to participants and related patient and public communities

No patients were involved in setting the research question or the outcome measures, nor were they involved in the design and implementation of the study. We plan to disseminate these findings to participants through the AP-HP website and to the general public in a press release.

