## Supplementary material for "Association between Hydroxyzine Use and Reduced Mortality in Patients Hospitalized for Coronavirus Disease 2019: Results from a multicenter observational study"

**eFigure 1. Course of the levels of the biological inflammatory markers LCRP (A), NLR (B) and IL-6 (C) during the visit according to hydroxyzine daily dose.** Vertical lines represent standard errors.

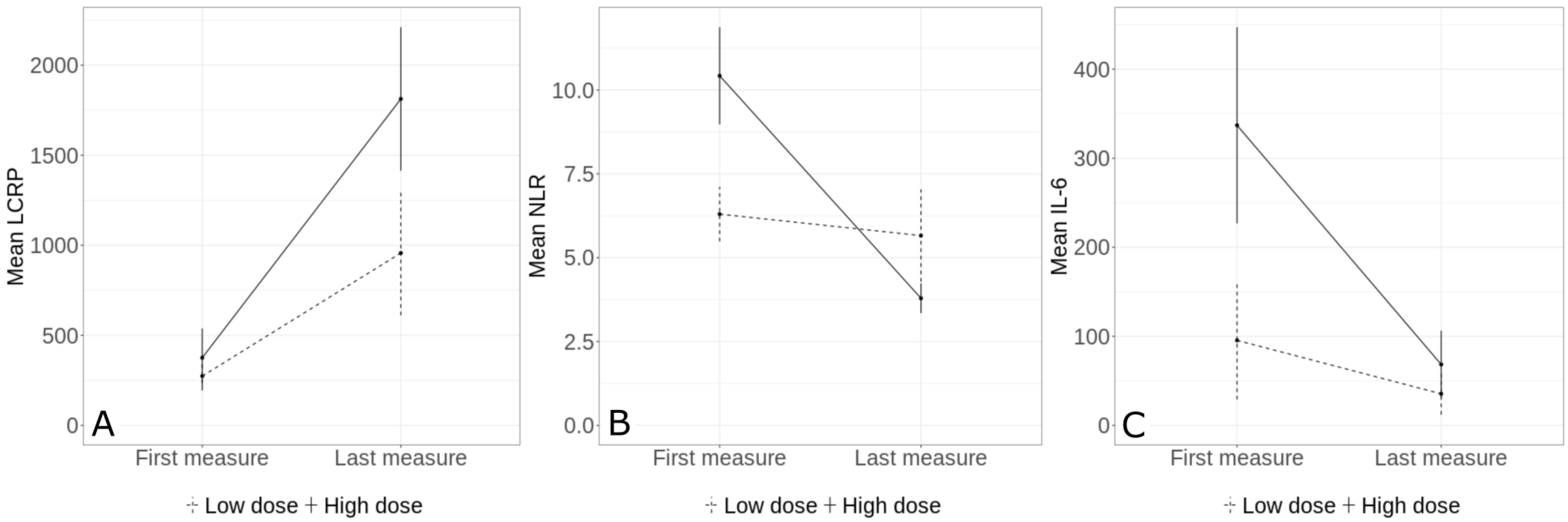

**eFigure 2. Quantitative bias analysis based on the observed residual differences in age (A) and other patients characteristics (B).**

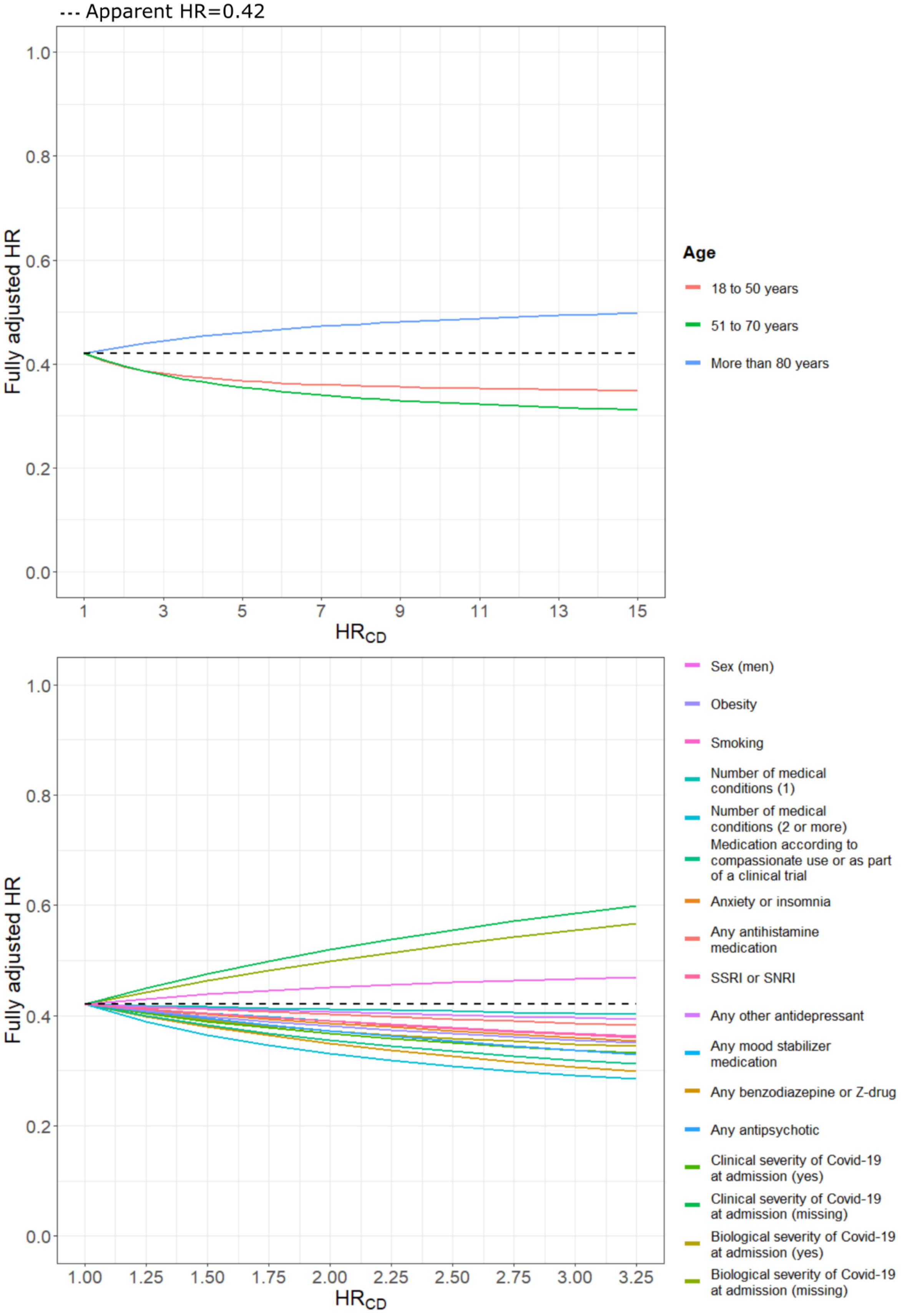

**eTable 1. Associations of baseline clinical characteristics with the endpoint of death in the cohort of patients who had been admitted to the hospital for Covid-19 (N=7,345).**

| | | | | Endpoint of death | | |
|---|---|---|---|---|---|---|
| | Full population (N=7,345) | With the end-point event (N=994) | Without the end-point event (N=6,351) | Crude analysis | Multivariable analysis (df=20) | |
| | N (%) | N (%) | N (%) | HR (SE) / p-value | HR (SE) / p-value | Collinearity diagnosis ($GVIF^{1/(2*df)}$) |
| Age | | | | | | 1.06 |
| *18 to 50 years* | 2709 (36.9%) | 42 (4.23%) | 2667 (42.0%) | Ref. | Ref. | |
| *51 to 70 years* | 2530 (34.4%) | 240 (24.1%) | 2290 (36.1%) | 5.31 (0.17) / <0.001* | 2.82 (0.20) / <0.001 | |
| *71 to 80 years* | 942 (12.8%) | 273 (27.5%) | 669 (10.5%) | 14.86 (0.17) / <0.001* | 7.53 (0.20) / <0.001 | |
| *More than 80 years* | 1164 (15.8%) | 439 (44.2%) | 725 (11.4%) | 16.99 (0.16) / <0.001* | 11.35 (0.19) / <0.001 | |
| Sex | | | | | | 1.06 |
| *Women* | 3619 (49.3%) | 359 (36.1%) | 3260 (51.3%) | Ref. | Ref. | |
| *Men* | 3726 (50.7%) | 635 (63.9%) | 3091 (48.7%) | 1.68 (0.07) / <0.001* | 1.32 (0.08) / <0.001 | |
| Obesity [α] | | | | | | 1.06 |
| *Yes* | 975 (13.3%) | 216 (21.7%) | 759 (12.0%) | 1.51 (0.08) / <0.001* | 1.23 (0.09) / 0.021 | |
| *No* | 6370 (86.7%) | 778 (78.3%) | 5592 (88.0%) | Ref. | Ref. | |
| Smoking [β] | | | | | | 1.03 |
| *Yes* | 623 (8.48%) | 149 (15.0%) | 474 (7.46%) | 1.51 (0.09) / <0.001* | 0.93 (0.10) / 0.452 | |
| *No* | 6722 (91.5%) | 845 (85.0%) | 5877 (92.5%) | Ref. | Ref. | |
| Number of medical conditions [γ] | | | | | | 1.12 |
| *0* | 4750 (64.7%) | 319 (32.1%) | 4431 (69.8%) | Ref. | Ref. | |
| *1* | 534 (7.27%) | 63 (6.34%) | 471 (7.42%) | 2.29 (0.14) / <0.001* | 1.78 (0.16) / <0.001 | |
| *2 or more* | 2061 (28.1%) | 612 (61.6%) | 1449 (22.8%) | 4.71 (0.07) / <0.001* | 3.49 (0.09) / <0.001 | |

| | | | | | | |
|---|---|---|---|---|---|---|
| Medication according to compassionate use or as part of a clinical trial Θ | | | | | | 1.11 |
| *Yes* | 1288 (17.5%) | 226 (22.7%) | 1062 (16.7%) | 1.13 (0.08) / 0.105 | 0.74 (0.09) / 0.001 | |
| *No* | 6057 (82.5%) | 768 (77.3%) | 5289 (83.3%) | Ref. | Ref. | |
| Anxiety or insomnia € | | | | | | 1.04 |
| *Yes* | 299 (4.07%) | 101 (10.2%) | 198 (3.12%) | 2.38 (0.11) / <0.001* | 1.24 (0.12) / 0.062 | |
| *No* | 7046 (95.9%) | 893 (89.8%) | 6153 (96.9%) | Ref. | Ref. | |
| Any other antihistamine medication | | | | | | 1.01 |
| *Yes* | 100 (1.4%) | 16 (1.6%) | 84 (1.3%) | 0.94 (0.25) / 0.799 | 0.67 (0.29) / 0.160 | |
| *No* | 7245 (98.6%) | 978 (98.4%) | 6267 (98.7%) | Ref. | Ref. | |
| SSRI or SNRI | | | | | | 1.08 |
| *Yes* | 318 (4.3%) | 81 (8.15%) | 237 (3.73%) | 1.5 (0.12) / <0.001* | 0.66 (0.12) / 0.001 | |
| *No* | 7027 (95.7%) | 913 (91.9%) | 6114 (96.3%) | Ref. | Ref. | |
| Any other antidepressant | | | | | | 1.05 |
| *Yes* | 142 (1.93%) | 47 (4.73%) | 95 (1.50%) | 2.03 (0.15) <0.001* | 0.85 (0.17) / 0.340 | |
| *No* | 7203 (98.1%) | 947 (95.3%) | 6256 (98.5%) | Ref. | Ref. | |
| Any mood stabilizer medication Ω | | | | | | 1.05 |
| *Yes* | 287 (3.91%) | 69 (6.94%) | 218 (3.43%) | 1.42 (0.12) / 0.005* | 0.93 (0.14) / 0.587 | |
| *No* | 7058 (96.1%) | 925 (93.1%) | 6133 (96.6%) | Ref. | Ref. | |
| Any benzodiazepine or Z-drug | | | | | | 1.14 |
| *Yes* | 752 (10.2%) | 268 (27.0%) | 484 (7.62%) | 2.53 (0.07) / <0.001* | 1.56 (0.09) / <0.001 | |
| *No* | 6593 (89.8%) | 726 (73.0%) | 5867 (92.4%) | Ref. | Ref. | |
| Any antipsychotic | | | | | | 1.07 |

| | | | | | | |
|---|---|---|---|---|---|---|
| *Yes* | 264 (3.59%) | 60 (6.04%) | 204 (3.21%) | 1.26 (0.13) / 0.086 | 0.80 (0.14) / 0.120 | |
| *No* | 7081 (96.4%) | 934 (94.0%) | 6147 (96.8%) | Ref. | Ref. | |
| Clinical severity of Covid-19 at admission [μ] | | | | | | 1.15 |
| *Yes* | 1564 (21.3%) | 421 (42.4%) | 1143 (18.0%) | 1.81 (0.08) / <0.001* | 1.69 (0.09) / <0.001 | |
| *No* | 1858 (25.3%) | 265 (26.7%) | 1593 (25.1%) | Ref. | Ref. | |
| *Missing* | 3923 (53.4%) | 308 (31.0%) | 3615 (56.9%) | 0.59 (0.08) / <0.001* | 1.66 (0.11) / <0.001 | |
| Biological severity of Covid-19 at admission [κ] | | | | | | 1.15 |
| *Yes* | 2439 (33.2%) | 592 (59.6%) | 1847 (29.1%) | 1.94 (0.08) / <0.001* | 1.51 (0.09) / <0.001 | |
| *No* | 1861 (25.3%) | 257 (25.9%) | 1604 (25.3%) | Ref. | Ref. | |
| *Missing* | 3045 (41.5%) | 145 (14.6%) | 2900 (45.7%) | 0.39 (0.1) / <0.001* | 0.75 (0.12) / 0.022 | |

[α] Defined as having a body-mass index higher than 30 kg/$m^2$ or an International Statistical Classification of Diseases and Related Health Problems (ICD-10) diagnosis code for obesity (E66.0, E66.1, E66.2, E66.8, E66.9).

[β] Smoking status was self-reported.

[γ] Assessed using ICD-10 diagnosis codes for diabetes mellitus (E11), diseases of the circulatory system (I00-I99), diseases of the respiratory system (J00-J99), neoplasms (C00-D49), diseases of the blood and blood-forming organs and certain disorders involving the immune mechanism (D5-D8), delirium (F05, R41) and dementia (G30, G31, F01-F03).

[ϴ] Any medication prescribed as part of a clinical trial or according to compassionate use (e.g., hydroxychloroquine, azithromycin, remdesivir, tocilizumab, sarilumab or dexamethasone).

[€] Assessed using ICD-10 diagnosis codes for anxiety, dissociative, stress-related, somatoform and other nonpsychotic mental disorders (F40-F48) and insomnia (G47).

[Ω] Included lithium or antiepileptic medications with mood stabilizing properties.

[μ] Defined as having at least one of the following criteria: respiratory rate > 24 breaths/min or < 12 breaths/min, resting peripheral capillary oxygen saturation in ambient air < 90%, temperature > 40°C, or systolic blood pressure < 100 mm Hg.

[κ] Defined as having at least one of the following criteria: high neutrophil-to-lymphocyte ratio, low lymphocyte-to-C-reactive protein (both variables were dichotomized at the median of the values observed in the full sample), and plasma lactate levels higher than 2 mmol/L.

* p-value is significant (p<0.05).

Abbreviations: HR, hazard ratio; SE, standard error; GVIF, generalized variance inflation factor.

**eTable 2. Association between hydroxyzine use within the 3 months prior to hospital admission and not during the visit for COVID-19 and the endpoint of death in the full sample and in the matched analytic sample.**

| | **Number of events / Number of patients** | **Crude Cox regression analysis** | **Multivariable Cox regression analysis** | **Analysis weighted by inverse-probability-weighting weights** | **Number of events / Number of patients** | **Univariate Cox regression in matched analytic samples** |
|---|---|---|---|---|---|---|
| | N (%) | HR (95% CI; p-value) | HR (95% CI; p-value) (df=23) | HR (95% CI; p-value) | N (%) | HR (95% CI; p-value) |
| Hydroxyzine use during the visit for COVID-19 | 12 / 125 (9.6%) | Ref. | Ref. | Ref. | 12 / 125 (9.6%) | Ref. |
| Hydroxyzine only within the last 3 months prior to the visit | 19 / 51 (37.3%) | 3.25 (1.58 - 6.71; 0.001*) | 3.43 (1.65 – 7.12; 0.001*) | 3.28 (1.54 – 6.98; 0.002*) | 19 / 51 (37.3%) | 3.02 (1.46 – 6.24; 0.003*) |
| Hydroxyzine use within the last 3 months and during the visit | 3 / 13 (23.1%) | 2.07 (0.59 – 7.36; 0.258) | 2.88 (0.80 – 10.30; 0.104) | 2.05 (0.57 – 7.46; 0.274) | 3 / 13 (23.1%) | 1.85 (0.52 – 6.59; 0.340) |
| No hydroxyzine | 960 / 7,156 (13.4%) | 1.84 (1.04 – 3.25; 0.036*) | 2.60 (1.47 – 4.63; 0.001*) | 3.02 (1.68 – 5.42; <0.001*) | 42 / 189 (22.2%) | 2.94 (1.55 – 5.59; 0.001*) |

* p-value is significant (p<0.05).

**eTable 3. Comparing the associations of hydroxyzine use and zopiclone use with the endpoint of death in the full sample and in the matched analytic sample**.

| | **Number of events / Number of patients** | **Crude Cox regression analysis** | **Multivariable Cox regression analysis** | **Analysis weighted by inverse-probability-weighting weights** | **Number of events / Number of patients** | **Univariate Cox regression in the matched analytic sample (1:1)** |
|---|---|---|---|---|---|---|
| | N (%) | HR (95% CI; p-value) | HR (95% CI; p-value) (df=24) | HR (95% CI; p-value) | N (%) | HR (95% CI; p-value) |
| Zopiclone | 46 / 151 (30.5%) | Ref. | Ref. | Ref. | 4 / 12 (33.3%) | Ref. |
| Hydroxyzine | 14 / 129 (10.9%) | 0.38 (0.21 - 0.70; 0.002*) | 0.39 (0.21 – 0.73; 0.003*) | 0.32 (0.15 – 0.70; 0.004*) | 14 / 129 (10.9%) | 0.27 (0.09 – 0.83; 0.022*) |
| Hydroxyzine and Zopiclone | 1 / 9 (1.1%) | 0.27 (0.04 - 2.00; 0.201) | 0.29 (0.04 – 2.43; 0.255) | 0.06 (0.01 – 0.49; 0.009*) | 1 / 9 (1.1%) | 0.19 (0.02 – 1.68; 0.135) |
| No hydroxyzine or Zopiclone | 933 / 7056 (13.2%) | 0.61 (0.0.45 - 0.82; 0.001*) | 0.85 (0.62 – 1.16; 0.305) | 0.74 (0.51 – 1.08; 0.116) | 49 / 264 (18.6%) | 0.61 (0.22 – 1.70; 0.349) |

* p-value is significant (p<0.05).

**eTable 4. Associations between hydroxyzine daily dose and the endpoint of death among patients using hydroxyzine during the visit (N=138)**.

| | Number of events / Number of patients | Crude Cox regression analysis | Cox regression adjusted for age and sex | Multivariable Cox regression analysis |
|---|---|---|---|---|
| | N (%) | HR (95% CI; p-value) | HR (95% CI; p-value) (df=6) | HR (95% CI; p-value) (df=22) |
| ***Daily dose*** | | | | |
| Lower doses | 8 / 53 (15.1%) | Ref. | Ref. | Ref. |
| Higher doses | 2 / 53 (3.8%) | 0.11 (0.02 – 0.48; 0.004*) | 0.12 (0.03 – 0.57; 0.007*) | 0.10 (0.02 – 0.45; 0.003*) |

* p-value is significant (p<0.05).
Because daily dose could not be ascertained with certainty in 32 (23.2%) patients, these patients have been excluded from this analysis.
The variable ‘daily dose’ has been dichotomized by the median.

**eTable 5. Association between levels of the biological inflammatory markers NLR, LCRP, IL-6 and lactates and the endpoint of death in the subsamples of patients with COVID-19 who had at least two measures of each marker during the visit.**

| | First measure | | Last measure | |
|---|---|---|---|---|
| | **Number of events / Number of patients** | **Crude Cox regression analysis** $^{\alpha}$ | **Number of events / Number of patients** | **Crude Cox regression analysis** $^{\alpha}$ |
| | Events / N (%) | HR (95% CI; p-value) | Events / N (%) | HR (95% CI; p-value) |
| **Biological inflammatory markers** | | | | |
| LCRP | 426 / 1,879 (22.7%) | 0.83 (0.76 - 0.90; <0.001*) | 423 / 1,876 (22.5%) | 0.47 (0.43 - 0.60; <0.001*) |
| NLR | 492 / 2,158 (22.8%) | 1.26 (1.11 - 1.43; <0.001*) | 493 / 2,159 (22.8%) | 2.71 (2.47 - 2.96; <0.001*) |
| IL-6 | 53 / 315 (16.8%) | 1.29 (1.06 - 1.57; 0.010*) | 53 / 3,15 (16.8%) | 1.53 (1.33 - 1.76; <0.001*) |
| Lactates | 319 / 1,159 (27.5%) | 1.27 (0.79 – 2.03; 0.326) | 823 / 1,163 (27.8%) | 2.75 (2.43 – 3.13; <0.001*) |

$^{\alpha}$ Values were log-transformed to address non-normal distribution of the biological variables.

**eTable 6. Course of the levels of the biological inflammatory markers NLR, LCRP, IL-6 and lactates during the visit according to hydroxyzine use.**

| | **Full population** | **Exposed to hydroxyzine** | **Not exposed to hydroxyzine** | **Two-way ANOVA: Interaction term** |
|---|---|---|---|---|
| | N / Mean (SD) | N / Mean (SD) | N / Mean (SD) | F (p) [α] |
| **Biological inflammatory markers** | | | | |
| LCRP | | | | 16.33 (<0.001*) |
| *First measurement* | 1,879 / 695.1 (2,732.0) | 63 / 308.4 (666.3) | 1,816 / 708.5 (2,775.3) | |
| *Last measurement* | 1,879 / 1,396.2 (3,831.7) | 63 / 1,640.5 (2,673.9) | 1,816 / 1,387.7 (3,865.9) | |
| NLR | | | | 14.80 (<0.001*) |
| *First measurement* | 2,161 / 7.1 (6.8) | 72 / 8.3 (6.8) | 2,089 / 7.1 (6.8) | |
| *Last measurement* | 2,161 / 6.8 (9.1) | 72 / 4.9 (5.8) | 2,089 / 6.8 (9.2) | |
| IL-6 | | | | 5.87 (0.016*) |
| *First measurement* | 315 / 182.4 (405.7) | 10/ 216.2 (230.1) | 305 / 181.2 (410.4) | |
| *Last measurement* | 315 / 238.1 (800.2) | 10 / 51.9 (68.6) | 305 / 244.2 (812.4) | |
| Lactates | | | | 0.94 (0.333) |
| *First measurement* | 1,142 / 1.5 (0.8) | 43 / 1.4 (0.7) | 1,099 / 1.5 (0.8) | |
| *Last measurement* | 1,142 / 1.5 (1.3) | 43 / 1.2 (0.5) | 1,099 / 1.6 (1.3) | |

[α] Values were log-transformed to address non-normal distribution of the biological variables.

* p-value is significant (p<0.05).

**eTable 7. Course of the levels of the biological inflammatory markers NLR, LCRP, and IL-6 during the visit according to hydroxyzine dose.**

| | Full population | Low dosage | High dosage | Two-way ANOVA: Interaction term |
|---|---|---|---|---|
| | N / Mean (SD) | N / Mean (SD) | N / Mean (SD) | F (p)$^{\alpha}$ |
| **Biological inflammatory markers** | | | | |
| LCRP | | | | 4.57 (0.037*) |
| *First measurement* | 55 / 329.2 (708.6) | 25 / 273.5 (399.4) | 30 / 375.6 (893.4) | |
| *Last measurement* | 55 / 1,423.8 (2,016.8) | 25 / 957.2 (1,728.3) | 30 / 1,812.7 (2,181.2) | |
| NLR | | | | 5.62 (0.021*) |
| *First measurement* | 60 / 8.6 (7.2) | 26 / 6.3 (4.1) | 34 / 10.4 (8.4) | |
| *Last measurement* | 60 / 4.6 (5.0) | 26 / 5.7 (7.0) | 34 / 3.8 (2.6) | |
| IL-6 | | | | 0.62 (0.454) |
| *First measurement* | 10 / 216.2 (230.1) | 5 / 95.5 (148.4) | 337.0 (246.3) | |
| *Last measurement* | 10 / 51.9 (68.6) | 5 / 35.5 (52.3) | 68.4 (84.7) | |

$^{\alpha}$ Values were log-transformed to address non-normal distribution of the biological variables.
* p-value is significant ($p<0.05$).
